# Comparison of the Immunogenicity of five COVID-19 vaccines in Sri Lanka

**DOI:** 10.1101/2021.12.15.21267834

**Authors:** Chandima Jeewandara, Inoka Sepali Aberathna, Saubhagya Danasekara, Laksiri Gomes, Suranga Fernando, Dinuka Guruge, Thushali Ranasinghe, Banuri Gunasekera, Achala Kamaladasa, Heshan Kuruppu, Gayasha Somathilake, Jeewantha Jayamali, Deshni Jayathilaka, Helanka Dinesh Kumara Wijayatilake, Pradeep Darshana Pushpakumara, Michael Harvie, Thashmi Nimasha, Shiromi Devika Grace de Silva, Ruwan Wijayamuni, Lisa Schimanski, Pramila Rijal, Jack Tan, Alain Townsend, Graham S. Ogg, Gathsaurie Neelika Malavige

**Affiliations:** Allergy Immunology and Cell Biology Unit, Department of Immunology and Molecular Medicine, University of Sri Jayewardenepura, Nugegoda, Sri Lanka; Ministry of Health, Sri Lanka; Colombo Municipal Council, Colombo, Sri Lanka; MRC Human Immunology Unit, MRC Weatherall Institute of Molecular Medicine, University of Oxford, Oxford, United Kingdom; Centre for Translational Immunology, Chinese Academy of Medical Sciences Oxford Institute, University of Oxford, Oxford, United Kingdom

## Abstract

We assessed antibody responses 3 months post-vaccination in those who received mRNA-1273 (n=225), Sputnik V (n=128) or the first dose of Gam-COVID-Vac (n=184) and compared the results with previously reported data of Sinopharm and AZD1222 vaccinees. 99.5% of Moderna >94% of AZD1222 or Sputnik V, 72% to 76% of Gam-COVID-Vac (first dose) and 38.1% to 68.3% of Sinopharm vaccinees had ACE2 blocking antibodies above the positive threshold. The ACE2 blocking antibody levels were highest to lowest was Moderna > Sputnik V/ AZD1222 (had equal levels)> first dose of Gam-COVID-Vac > Sinopharm. All Moderna recipients had antibodies above the positive threshold to the ancestral (WT), B.1.1.7, B.1.351.1 and 80% positivity rate for B.1.617.2. Positivity rates of Sputnik V vaccinees for WT and variants, were higher than AZD1222 vaccinees, while Sinopharm vaccinees had the lowest positivity rates (<16.7%). These findings highlight the need for further studies to understand the effects on clinical outcomes.

## Introduction

With the emergence and rapid spread of the Omicron variant, many high income and upper middle income countries have ramped up their vaccination programs by rolling out booster doses to all individuals over 18 years of age^1^, while many individuals in lower income countries are yet to receive their first dose^2^. There are currently seven COVID-19 vaccines which the WHO has given emergency use authorization^3^, while some other vaccines such as Gam-COVID-Vac (Sputnik V and Sputnik light) have been widely used without WHO emergency use license^4^. US, Europe and other high-income countries have vaccinated their populations largely either using an mRNA vaccine (mRNA-1273 or BNT162b2) or AZD1222, while many lower middle income countries and low income countries have been using inactivated vaccines such as Sinovac, Sinopharm (BBIBP-CorV) or the adenovirus vector vaccine Gam-COVID-Vac^4^.

The efficacy of the different COVID-19 vaccines varies widely, and the levels of neutralizing antibodies (Nabs) elicited by different vaccines have shown to correlate with efficacy rates^5^. A direct comparison between four different vaccines, between two to three months post immunization showed that the Pfizer-BioNTech (BNT162b2), elicited the highest ACE2 blocking antibodies, followed by AZD1222, Sputnik V and Sinopharm^6^. Furthermore, the waning of Nabs and T cell responses with time has been shown to vary widely for different vaccines^7-9^. These differences in the induction of Nabs and their persistence is likely to have a significant impact of the transmission dynamics of the SARS-CoV-2 variants of concern (VOC), especially with the emergence of Omicron. It was shown that a 41-fold decline in Nabs elicited by the Pfizer-BioNtech vaccine and 30 to 60 fold reduction of Nabs in convalescent plasma was observed for the Omicron variant^10,11^. This immune escape by Omicron was found to be less in those who were previously infected and vaccinated (ref). In order to prepare for the rapid spread of Omicron globally, many high-income countries have reduced the gap between the 2^nd^ dose and the booster to three months and to give a booster dose to all adults over 18 years of age^12,13^. Although it appears that the Omicron variant significantly evades immunity, induction of higher Nabs through giving a booster dose, is likely to reduce this immune escape (ref). However, the Nabs levels following booster doses would depend on the Nabs levels post-second dose. Furthermore, although high-income countries are rapidly deploying booster doses and therefore could possibly reduce the impact due to the rapid transmission of Omicron, the transmission dynamics and clinical disease severity could be different in many lower middle-income countries with lower infection rates, and lower vaccination rates.

Sri Lanka has currently fully vaccinated 64% of its population, while 74% have received at least a single dose of the vaccine^4^. Sinopharm (BBIBP-CorV) was the main vaccine used with 11.9 million individuals^14^, which is 56.9% of the total population being vaccinated with this vaccine. Some individuals were also vaccinated with Sputnik V and due to the late arrival of the second dose of Gam-COVID-Vac, many individuals only received the first dose of the Gam-COVID-Vac (rAd26-S) for 3 months, which is marketed as a single dose vaccine. We had previously published the kinetics of antibody and T cell responses to the AZD1222 and the Sinopharm vaccine in the Sri Lankan population separately, which showed significant differences^8,9^. In order to get a better idea regarding the differences in immunogenicity of different vaccines, we wished to build on that data by carrying out a new analysis by carrying out a direct comparison for the immunogenicity of different vaccines, at the same time point post-vaccination. Therefore, we sought to compare the antibody levels to the receptor binding domain (RBD) of the SARS-CoV-virus, VOCs and ACE2 blocking antibodies, 3 months post vaccination, in Sri Lankan individuals who received two doses of Moderna (mRNA-1273), AZD1222, Sinopharm, Sputnik V or a single dose of Gam-COVID-Vac. We further compared the antibody levels in vaccinees who were uninfected and who were naturally infected to determine the changes in antibody levels with natural infection.

## Results

### Total antibody responses to the SARS-CoV2 receptor binding domain (RBD)

The number of individuals included in assessing total antibody responses (IgG, IgA and IgM) to the RBD of the virus is shown in table 1. As we have previously shown, all those who received the two doses of the AZ vaccine had seroconverted ^8^, whereas the overall seroconversion rate for Sinopharm was 95.07% ^9^. All individuals (100%) who had received both doses of the Sputnik vaccines had also seroconverted, whereas 43/45 (95.5%) of those aged 20 to 39 and 130/139 (93.5%) individuals aged 40 to 59 who received only the first dose of Sputnik, seroconverted. Of those who had received Moderna, 46/48 (95.8%) aged 20 to 39, 124/132 (93.9%) aged 40 to 59 and 41/44 (93.2%) individuals >60 were found to have seroconverted by this assay.

**Table 1:** The positivity rates for the WT and SARS-CoV-2 variants of concern, in different age groups in those who received two doses of Moderna or two doses of Sputnik V measured by the haemagglutination assay.

| Age groups | WT<br>N (%) | B.1.1.7<br>(alpha)<br>N (%) | B.1.351<br>(beta)<br>N (%) | B.1.617.2<br>(delta)<br>N (%) |
| --- | --- | --- | --- | --- |
| Moderna |  |  |  |  |
| 20 to 39 (n=26) | 26 (100 %) | 26 (100 %) | 26 (100 %) | 22 (84.62%) |
| 40 to 59 (n=25) | 25 (100 %) | 25 (100 %) | 25 (100 %) | 21 (84%) |
| >60 (n=13) | 13 (100 %) | 13 (100 %) | 13 (100 %) | 13 (100 %) |
| Sputnik V |  |  |  |  |
| 20 to 39 (n=25) | 20 (80%) | 17 (68%) | 17 (68%) | 19 (76%) |
| 40 to 59 (n=46) | 37 (80.43%) | 33 (71.74%) | 31 (67.39%) | 34 (73.91%) |

In individuals in the 20 to 39 age group, those who received 2 doses of AZD1222 and two doses of Sputnik V had significantly higher (p<0.0001) total antibody responses to the RBD than those who received two doses of the Sinopharm vaccine (Figure 1A). Those who received two doses of AZD1222 and both doses of Sputnik V also had significantly higher total antibody responses to the RBD than those who received only the first dose of Sputnik (p<0.0001) and both doses of Moderna (p<0.0001). Those who received both doses of Moderna and one dose of Sputnik had similar levels of total antibodies to the RBD as those who had both doses of Sinopharm (Figure 1A).

**Figure 1:**
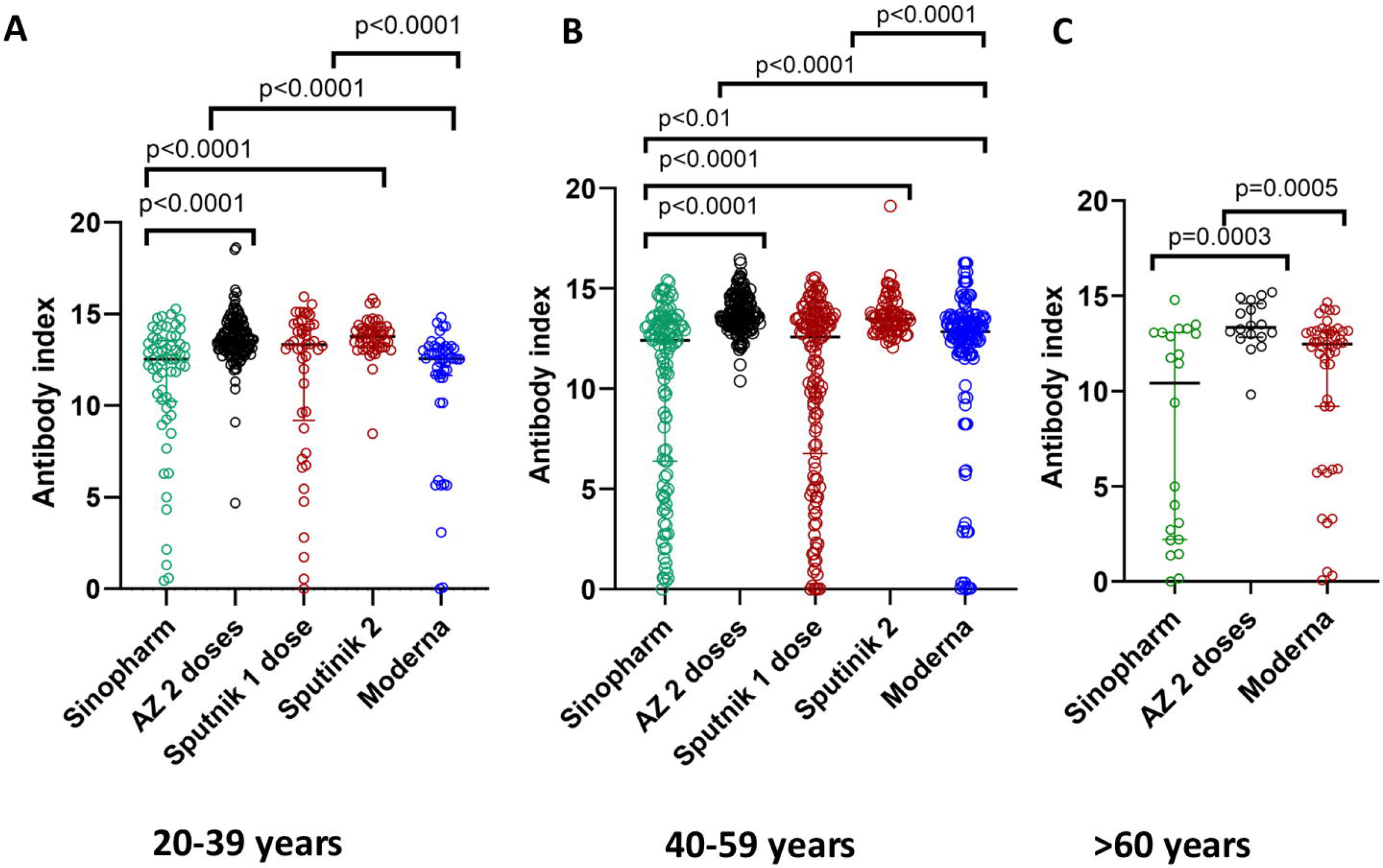
SARS-CoV-2 specific total antibodies to the receptor binding domain (RBD) in those who received different vaccines at 3 months following full vaccination. Total antibodies to the RBD were measured by ELISA in 20- to 39-year-olds who received two doses of Sinopharm (n=61), AZD1222 (120), Sputnik 1 dose (n=45), Sputnik 2 doses (n=50) and Moderna 2 doses (n=48) (A). Total antibodies to the RBD were also measured in 40 to 59-year-olds who received two doses of Sinopharm (n=120), AZD1222 (153), Sputnik 1 dose (n=139), Sputnik 2 doses (n=77) and Moderna 2 doses (n=132) (B). In those >60 years of age, the analysis was carried out in those who received 2 doses of Sinopharm (n=22), 2 doses of AZD1222 (n=18) and 2 doses of Moderna (n=24). The differences in antibody titres (antibody index) between different vaccines were analysed using the Mann-Whitney test. All tests were two-tailed. The lines indicate the median and the inter quartile range.

In individuals in the 40 to 59 age group, those who received 2 doses of AZD1222 and two doses of Sputnik V had significantly higher (p<0.0001) total antibody responses to the RBD than those who received two doses of the Sinopharm vaccine (Figure 1B). Individuals who had both doses of these vaccines also had significantly higher (p<0.0001) total antibody responses to the RBD than those who received one dose of Sputnik and both doses of Moderna. Those who had both doses of Moderna had significantly higher responses (p=0.01) than those who received both doses of Sinopharm (Figure 1B).

In those who were >60 years of age, the analysis was only carried out for Sinopharm, AZD1222 and Moderna as we had not recruited individuals >60 years who had received Sputnik vaccines. Those who had received 2 doses of AZD1222 had significantly higher total antibody responses to the RBD than those who had received both doses of Sinopharm (p=0.0003) and both doses of Moderna (p=0.0005) (Figure 1C). There was no significant difference between total antibody levels to the RBD in those who had received Sinopharm compared to Moderna (p=0.09).

### ACE2 blocking antibodies assessed by the surrogate neutralizing antibody test (sVNT) for different vaccines

ACE2 blocking antibodies were assessed in a sub cohort of uninfected individuals for each vaccine in each age group. The number of individuals included in each age group for each vaccine is shown in table 1. In the 20 to 39 age group, as previously reported ^9^, only 68.3% who received Sinopharm gave a positive result for the presence ACE2 blocking antibodies, while 96.2% of individuals who received two doses of AZD1222 gave a positive response ^8^. 19/25 (76%) of those who received only the first dose of Gam-COVID-Vac gave a positive response, while 125/128 (97.7%) of those who received two doses of Sputnik and 224/225 (99.5%) two doses of Moderna gave a positive response. Those who received two doses of AZD1222, Sputnik V and Moderna had significantly higher (p<0.0001) ACE2 blocking antibodies than those who received two doses of Sinopharm or one dose of Sputnik (Figure 2A). In contrast to what was observed with the SARS-CoV-2 total antibodies to the RBD, those who received two doses of Moderna had significantly higher (p<0.0001) ACE2 blocking antibodies than those who had two doses of AZD1222 or Sputnik V (Figure 2A). The median ACE2 blocking antibody levels for Moderna was 99.2% (IQR 98.8 to 99.4% of inhibition), while levels following two doses of Sputnik V was 88.2 (IQR 73.1 to 98.1 % of inhibition) and one dose of Gam-COVID-Vac was 48.1 (IQR 26.9 to 64.2%). The levels for AZD1222 were 85.2 (IQR 58.9 to 96.5% of inhibition) and for Sinopharm 37.7 (IQR 19.6 to 58.9% of inhibition) as previously reported^8,9^.

**Figure 2:**
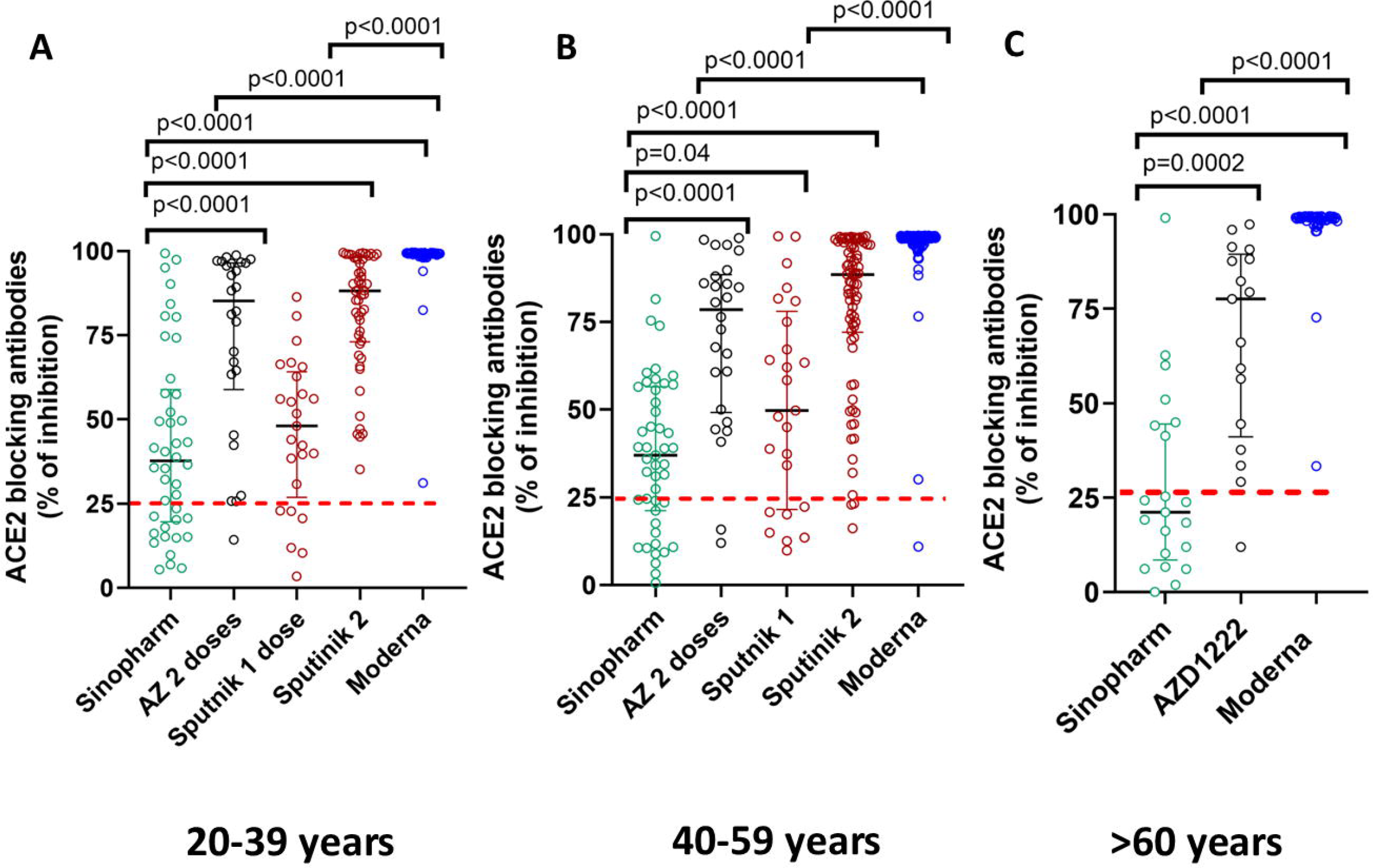
SARS-CoV-2 ACE2 blocking antibodies in those who received different vaccines at 3 months following full vaccination. ACE2 blocking antibodies were measured by the sVNT assay in 20 to 39 year olds who received two doses of Sinopharm (n=41), AZD1222 (26), Sputnik 1 dose (n=25), Sputnik 2 doses (n=50) and Moderna 2 doses (n=48) (A). ACE2 blocking antibodies were also measured in 40 to 59-year-olds who received two doses of Sinopharm (n=48), AZD1222 (36), Sputnik 1 dose (n=25), Sputnik 2 doses (n=77) and Moderna 2 doses (n=132) (B). In those >60 years of age, the analysis was carried out in those who received 2 doses of Sinopharm (n=21), 2 doses of AZD1222 (n=17) and 2 doses of Moderna (n=44). The differences in ACE2 blocking antibodies (% of inhibition) between different vaccines were analysed using the Mann-Whitney test. All tests were two-tailed. The lines indicate the median and the inter quartile range. The positive cut-off value if shown as a red dotted line.

In the 40 to 59 age group, as reported before, 66.7% of those who received two doses of Sinopharm ^9^ and 94.5% of those who received two doses of AZD1222 ^8^ gave a positive response. 18/25 (72%) of those who received only one dose of Gam-COVID-Vac and 74/77 (96.1%) of those who received both doses of Sputnik V and 135/136 (99.4%) who received both doses of Moderna gave a positive result. Again, those who received two doses of AZD1222, Sputnik V and Moderna had significantly higher (p<0.0001) ACE2 blocking antibodies than those who received two doses of Sinopharm or one dose of Gam-COVID-Vac (Figure 2B). Those who received one dose of Gam-COVID-Vac also had significantly higher levels (p=0.04) of antibodies than those who received two doses of Sinopharm. Again, those who received two doses of Moderna had significantly higher (p<0.0001) ACE2 blocking antibodies than those who had two doses of AZD1222 or Sputnik V (Figure 2B). The median ACE2 blocking antibody levels for Moderna was 98.9% of inhibition (IQR 98.9 to 99.3% of inhibition), while levels following two doses of Sputnik V was 88.4 (IQR 72.0 to 98.6 % of inhibition) and one dose of Gam-COVID-Vac was 49.7 (IQR 21.4 to 78.0%). The levels for AZD1222 were 78.5% inhibition (IQR 49.1 to 88.4 % of inhibition) and for Sinopharm 37.0% (IQR 21.2 to 56.5% of inhibition) as previously shown by us^8,9^.

As we could not recruit those >60 years of age to study the immunogenicity of the Sputnik vaccines. The analysis was limited to those who received Sinopharm, AZD1222 and Moderna. We had reported the results of those who received Sinopharm and AZD1222 in >60 years previously ^8,9^, which showed 38.1% of those who received Sinopharm and 94.1% of those who received AZD1222 gave a positive response. All individuals (100%) who were >60 and received Moderna both doses (n=44) gave a positive response for the presence of ACE2 blocking antibodies. The ACE2 blocking antibodies were significantly higher in those who received 2 doses of AZD1222 (p=0.0002) and Moderna (p<0.0001) compared to those who received Sinopharm (Figure 2C). The ACE2 blocking antibody levels were also significantly higher (p<0.0001) in those who received Moderna compared to those who received AZD1222 (Figure 2C). The median ACE2 blocking antibody levels for Moderna was 99.0% (IQR 98.2 to 99.4% of inhibition), while the levels for AZD1222 were 77.6 (IQR 41.0 to 89.4 % of inhibition) and for Sinopharm 21.1 (IQR 8.4 to 44.5% of inhibition) as previously shown by us^8,9^.

### SARS-CoV-2 RBD specific antibodies measured by the haemagglutination assay (HAT) for the ancestral virus and the VOCs

Our results showed that those who were given two doses of Moderna had significantly higher ACE2 blocking antibodies than those who received the two adenovirus vector vaccines, AZD1222 and Sputnik V. Therefore, we proceeded to investigate the differences in the antibody responses to RBD of the SARS-CoV-2 and VOCs by using the HAT assay, as this assay too has shown to correlate with neutralizing antibodies ^15^. The positivity rates of those who received two doses of Moderna and two doses of Sputnik V in different age groups is shown in table 1. Similar to the results seen with the sVNT assay, all those who received two doses of Moderna had a positive response to the SARS-CoV-2 ancestral strain (WT), alpha and beta variants, whereas positivity rates were 84% in the 20 to 39 and 40 to 59 age groups for delta. In contrast, the positivity rates for the WT and VOC in those who received two doses of Sputnik V were between 67% and 80%. Although these positivity rates were higher than those seen following two doses of AZD1222 (50 to 65%) in 20 to 39 and 40 to 59 year age groups^8^, the positivity rates were less than those following Moderna.

The HAT titres for the WT were significantly higher following Moderna compared to both doses of Sputnik V in 20 to 39 and 40 to 59 age groups (p<0.0001) (Figure 3A). HAT titres were also significantly higher for B.1.1.7 in the 20 to 39 (p=0.03) and the 40 to 59 (p=0.02) age groups for Moderna compared to Sputnik V although the difference was less than for the WT (Figure 3B). For B.1.315, a significant difference between HAT titres was only seen in the 40 to 59 age group, with those who received Moderna having significantly higher (p=0.002) levels (Figure 3C) whereas no difference was seen in the HAT titre levels for B.1.617.2 between either vaccine for any of the two age groups (Figure 3D).

**Figure 3:**
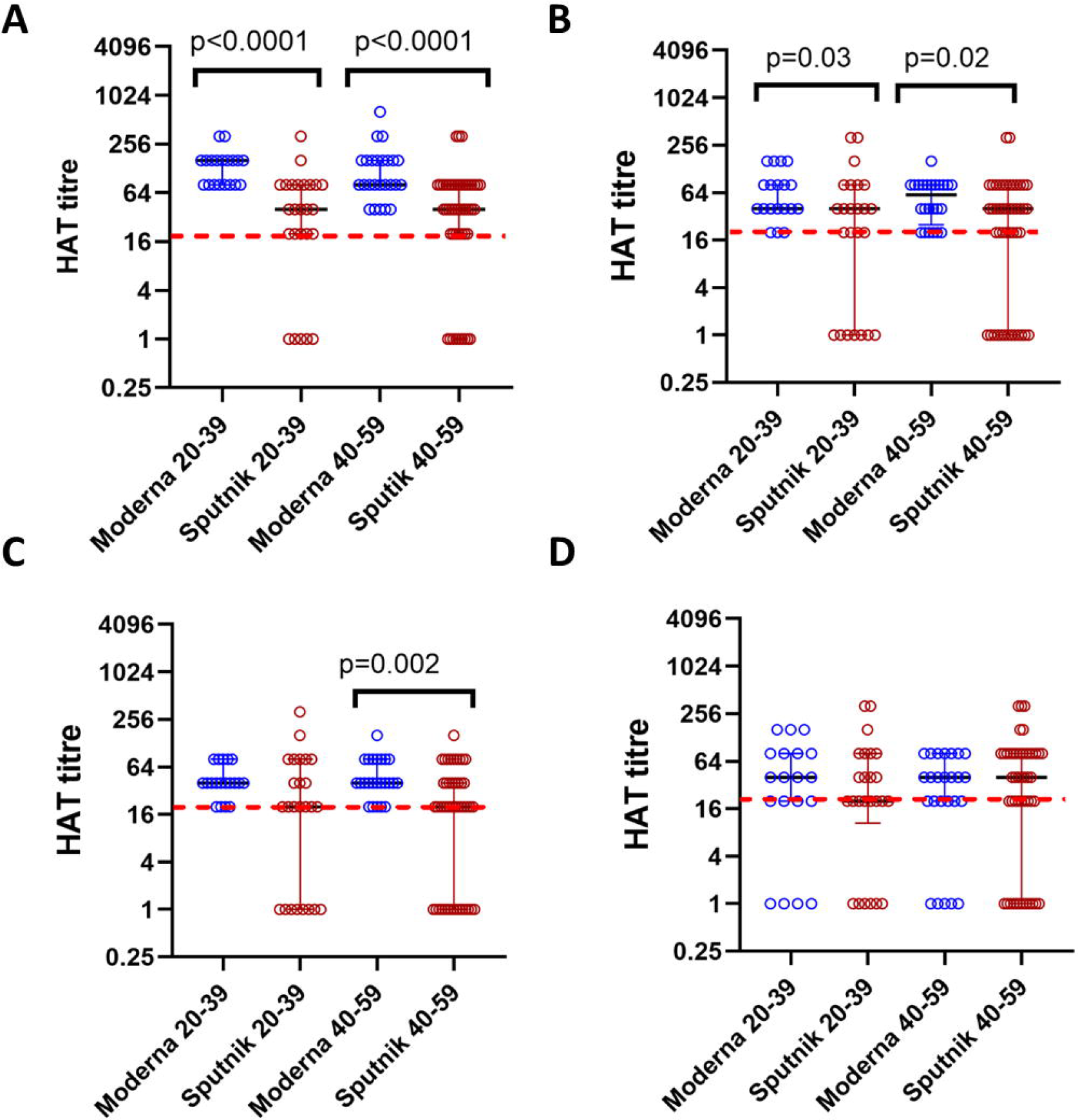
SARS-CoV-2 specific antibodies to the RBD of the ancestral (WT) virus and VOCs in those who received Spuntnik V and Moderna 3 months following the second dose. Antibodies to the RBD were measured by the haemagglutination test (HAT) in those who received two doses of Moderna in 20 to 39 years old (n=26) or two doses of Sputnik V (n=25) and those who were aged 40 to 59 years who received two doses of Moderna (n=25) or Sputnik V (n=46). Antibodies were measured by HAT to the WT (A), B.1.1.7 (B), B.1.351.1 (C) and B.1.617.2 (D). The differences between HAT titres for between the two vaccines were analysed using the Mann-Whitney test. All tests were two-tailed. The lines indicate the median and the inter quartile range. The positive cut-off value if shown as a red dotted line.

### SARS-CoV-2 total antibody responses and ACE2 blocking antibodies in those who were naturally infected and vaccinated

Individuals with previous natural infection are advised to obtain both doses of a COVID-19 vaccine in some countries, while only one dose of a vaccine is given in many countries in Europe. Therefore, in order to determine the immune responses in those who had natural infection and the different vaccines, we compared to immune responses of those who were fully vaccinated with one of the COVID-19 vaccines used in Sri Lanka (those who received the first dose of Sputnik were also investigated separately). The SARS-CoV-2 total antibodies and the ACE2 blocking antibody positivity and levels are shown in table 2. As the number of infected individuals who received different vaccines were small, we did not analyses the antibody levels for these different assays age groups.

**Table 2.**
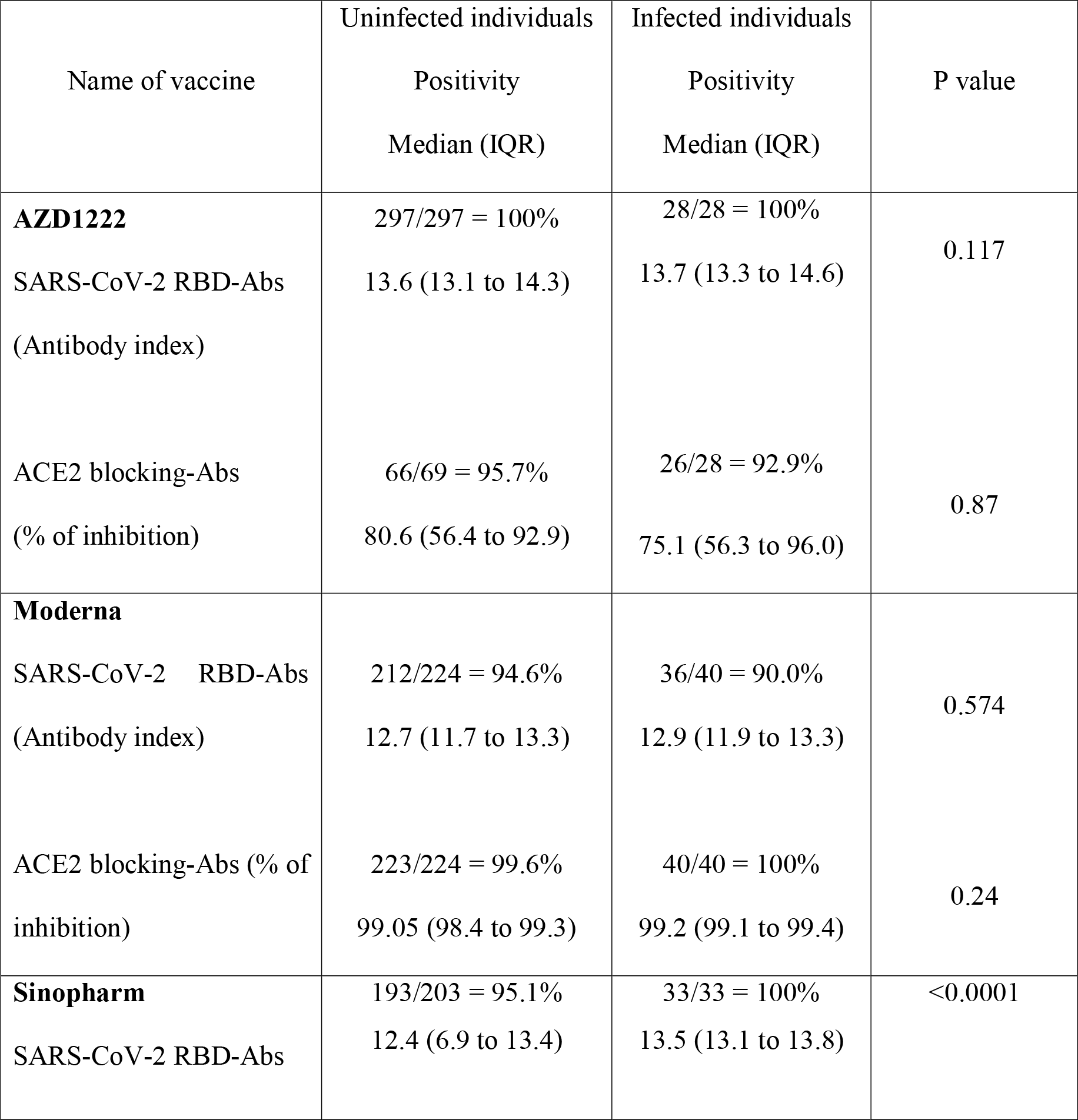
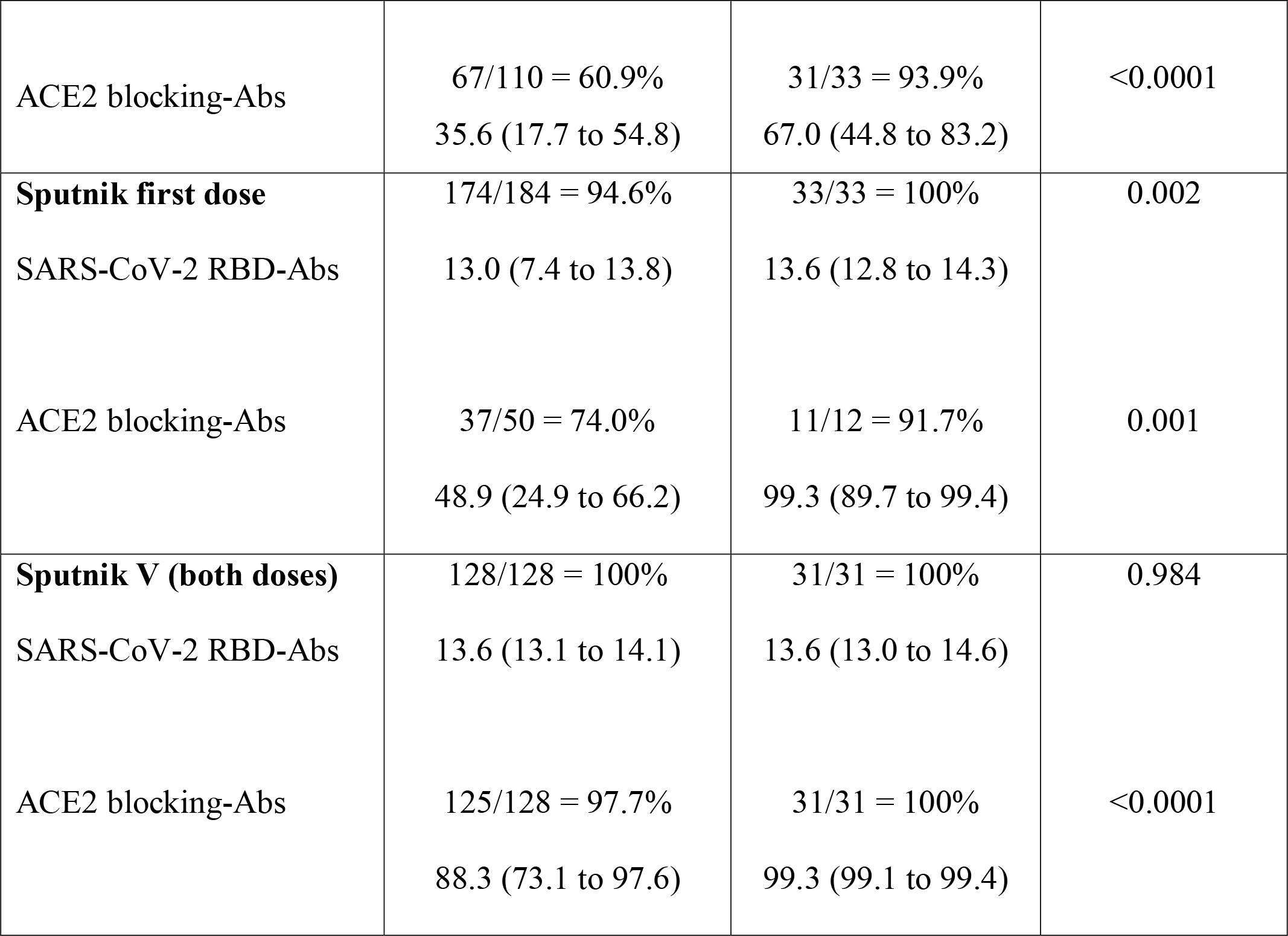
The positivity rates and median antibody titres for the RBD of SARS-CoV-2 (total antibodies) and ACE2 blocking antibodies in uninfected and infected vaccine recipients.

As shown in table 3, the seropositivity rates for the RBD of the SARS-CoV-2 (total antibody levels) and ACE2 blocking antibodies were significantly higher in infected individuals 3 months post-immunization, in those who received two doses of Sinopharm or one dose of Sputnik. For those who received two doses of Moderna, AZD1222 or Sputnik V, there was no difference in the total antibody levels to the RBD. However, the ACE2 blocking antibodies were significantly higher in infected individuals following two doses of Sputnik V vaccines, compared to uninfected vaccinees, whereas no difference was seen between uninfected and infected individuals who received AZD1222 or Moderna.

## Discussion

In this study we have compared the total antibody levels to the RBD of the SARS-CoV-2 virus, antibody levels to the RBD of VOCs and ACE2 blocking antibodies in those who received two doses of AZD1222, Moderna (mRNA-1273), Sinopharm or Sputnik V or the first dose of Gam-COVID-Vac (Sputnik light), 3 months post-immunization analysed at a single centre in Sri Lanka based on published and new analyses^8,9^. We found that 99.5% of those, in all age groups who received two doses of Moderna had a positive response for the presence of ACE2 blocking antibodies, which is a surrogate measure for the presence of neutralizing antibodies (Nabs), whereas the positivity rates for those who received two doses of AZD1222 or Spuntik V was over 94%. In contrast, the positivity rates following the first dose of Gam-COVID-Vac was 72% to 76% and as we previously reported for Sinopharm it ranged between 38.1% to 68.3% in different age groups^9^. Those who received Moderna also had significantly higher levels of ACE2 blocking antibodies than those who were given other vaccines. While the ACE2 blocking antibody levels following two doses of AZD1222 or Sputnik V were comparable, they were significantly higher than those who were given only one dose of Gam-COVID-Vac or two doses of Sinopharm. Nab levels have shown to strongly correlate with the level of protection against symptomatic COVID-19^5^ but the emergence of variants highlights the relevance of immunity beyond spike.

Nab levels have also shown to be a correlate of vaccine efficacy and booster doses were shown to increase the vaccine efficacy by increasing Nab titres^16^. The surrogate Nab test (sVNT) that measures ACE2 blocking antibodies has been widely used as a surrogate measure for Nabs^17-19^. While it has been shown that Nabs for current COVID-19 vaccines vary by as much as 25-fold, our data show that there are significant differences in the persistence of Nabs 3 months post-vaccination in different age groups that received different vaccines. It has been shown that waning of immunity following two doses of Moderna and AZD1222 associates with break-through infections, including an increase in hospitalization rates, especially after 20 weeks^20^ although waning of efficacy was less following Moderna^21^. The reduction in break-through infections and hospitalizations correlated with the increase in Nabs following the booster doses^22,23^. Although there are no data regarding the effectiveness of Sinopharm, one dose of Gam-COVID-Vac and Spuntik V in preventing breakthrough infections, hospitalizations and severe disease, based on the Nabs derived from the sVNT assay, those who received Sinopharm and one dose of Gam-COVID-Vac had substantially less ACE2 blocking antibodies than those who received Moderna, AZD1222 or two doses of Sputnik V. Sinopharm is an inactivated vaccine and so induces immune responses beyond spike which may be relevant, and were not tested here. We have not analysed T cell responses which may also impact, but overall the differences detected here are likely to be relevant with the emergence of the Omicron variant, which has a potential to further evade immunity^24^.

In this study we found that although the mRNA-1273 induced the highest levels of ACE2 blocking antibodies in all age groups and all vaccinees had levels above the cut-off value, the total antibody levels measured by the commercial Wantai antibody assay, which detects IgM, IgG and IgA antibodies to the RBD of the virus was significantly lower compared to those who received either two doses of AZD1222 or Sputnik V. In fact, 4.2% to 12.5% of those who received the Moderna vaccine did not have detectable antibodies to the RBD, whereas those who received two doses of the adenovirus vector vaccines had similar levels and significantly higher levels, with all individuals being seropositive by this assay. Since the findings based on this assay were different to those of the sVNT assay, we compared the antibodies to the RBD of the WT and VOCs by the haemagglutination assay (HAT), which was shown to correlate with the Nabs and with the sVNT, in those who received two doses of Moderna or Sputnik V ^15,25^. With the HAT assay again, all (100%) those who received Moderna had a positive response to WT, B.1.1.7 and B.1.351.1 and 84 to 100% to B.1.617.2. The positivity rates in those who received both doses of Sputnik V were between 67.4% to 80%, with titres significantly lower than those who received Moderna, although the positivity rates were higher than those who received two doses of AZD1222^8^. Since the HAT assay was shown to strongly correlate with Nabs levels^15^, it appears that while those who received Moderna had significantly higher Nabs than those who received other vaccines, those who received the two adenovirus vector vaccines had higher antibody levels to the RBD. While the reasons for these differences are not clear, it could be due to mRNA vaccines having stabilizing substitutions in spike protein to maintain the pre-fusion conformation, whereas AZD1222 and Sputnik may not contain these specific substitutions ^26,27^. However, it is possible that these differences are seen due to cross reactive antibody responses to different VOCs elicited by different vaccines. While the sVNT that detects ACE2 blocking antibodies would predominantly detect antibodies to the RBD of the WT, the HAT assay could be picking up antibodies that are cross reactive to other VOCs^28^. Therefore, it would be important to conduct a prospective study to understand if these differences in antibody levels and positivity rates observed with different assays translate to risk of infection or clinical disease severity.

With the emergence of Omicron many high income countries have now focused their vaccination programs in rapidly rolling out booster doses^1^. While some countries only gave one dose of a COVID-19 vaccine to those who had been previously had COVID-19^29^, there has not been guidance by many authorities in the need of booster doses for those who were fully vaccinated and infected. Our data show that in those who were infected and received two doses of either the Moderna or AZD1222, there was no difference in the ACE2 blocking antibodies in infected individuals compared to those who were uninfected, whereas for other vaccines the ACE2 blocking antibodies were significantly higher in those who were infected. The ACE2 blocking antibodies were over 99% in those who received one dose of Gam-COVID-Vac, Spuntik V or Moderna, while the median ACE2 blocking antibody levels were 67% and 75.1% for those who received two doses of Sinopharm or AZD1222 respectively. Therefore, infected individuals who received these vaccines could benefit from receiving a booster dose of the vaccine.

In summary, we have investigated antibody levels to the RBD of the ancestral SARS-CoV-2 virus, VOCs and ACE2 blocking antibodies in those who received five types of vaccines, 3 months post-vaccination in Sri Lanka based on published and new analyses from a single centre. We found that the seropositivity rates, ACE2 blocking antibody levels and antibodies to the RBD of VOCs showed a huge variation between vaccines. The levels of ACE2 blocking antibodies at 3 months post vaccination was highest was for Moderna, with Sputnik V and AZD1222 eliciting equal levels, followed by those who received the first dose of Gam-COVID-Vac (Sputnik light) and then Sinopharm. These differences in the persistence of immunity to different vaccines is likely to have significant implications in breakthrough infection rates, hospitalization and severe disease in different vaccine recipients.

## Supporting information

Detailed methods

## Data Availability

All data produced in the present work are contained in the manuscript.

## Funding statement

We are grateful to the Allergy, Immunology and Cell Biology Unit, University of Sri Jayewardenepura; the NIH, USA (grant number 5U01AI151788-02), UK Medical Research Council and the Foreign and Commonwealth Office for support. T.K.T. is funded by the Townsend-Jeantet Charitable Trust (charity number 1011770) and the EPA Cephalosporin Early Career Researcher Fund. A.T. are funded by the Chinese Academy of Medical Sciences (CAMS) Innovation Fund for Medical Science (CIFMS), China (grant no. 2018-I2M-2-002).

## Notes

### Competing Interest Statement

The authors have declared no competing interest.

### Author Declarations

Ethics approval was obtained from the Ethics Review Committee of University of Sri Jayewardenepura.

## References

1 Dolgin, E. Omicron is supercharging the COVID vaccine booster debate. Nature, doi:10.1038/d41586-021-03592-2 (2021).

2 Dong, E., Du, H. & Gardner, L. An interactive web-based dashboard to track COVID-19 in real time. The Lancet infectious diseases 20, 533–534, doi:10.1016/S1473-3099(20)30120-1 (2020).

3 WHO. COVID-19 vaccine tracker and landscape. (World Health Organization, 2021).

4 Hannah Ritchie, E. O.-O., Diana Beltekian, Edouard Mathieu, Joe Hasell, Bobbie Macdonald, Charlie Giattino, Cameron Appel and Max Roser. Coronavirus (COVID-19) Vaccinations, <https://ourworldindata.org/covid-vaccinations> (2021).

5 Khoury, D. S. et al. Neutralizing antibody levels are highly predictive of immune protection from symptomatic SARS-CoV-2 infection. Nature medicine 27, 1205–1211, doi:10.1038/s41591-021-01377-8 (2021).

6 Dashdorj, N. J. et al. Direct comparison of antibody responses to four SARS-CoV-2 vaccines in Mongolia. Cell host & microbe, doi:10.1016/j.chom.2021.11.004 (2021).

7 Collier, A. Y. et al. Differential Kinetics of Immune Responses Elicited by Covid-19 Vaccines. The New England journal of medicine 385, 2010–2012, doi:10.1056/NEJMc2115596 (2021).

8 Jeewandara, C. et al. Kinetics of immune responses to the AZD1222/Covishield vaccine with varying dose intervals in Sri Lankan individuals. medRxiv, doi:10.1101/2021.10.27.21265561 (2021).

9 Jeewandara, C. et al. Persistence of antibody and T cell responses to the Sinopharm/BBIBP-CorV vaccine in Sri Lankan individuals. medRxiv, 2021.2010.2014.21265030, doi:10.1101/2021.10.14.21265030 (2021).

10 Schmidt, F. et al. Plasma neutralization properties of the SARS-CoV-2 Omicron variant. medRxiv, 2021.2012.2012.21267646, doi:10.1101/2021.12.12.21267646 (2021).

11 Cele, S. et al. SARS-CoV-2 Omicron has extensive but incomplete escape of Pfizer BNT162b2 elicited neutralization and requires ACE2 for infection. medRxiv, 2021.2012.2008.21267417, doi:10.1101/2021.12.08.21267417 (2021).

12 Services, U. S. D. o. H. H. COVID-19 Vaccine Booster Shots. (2021).

13 Department of Health and Social Care, U. (2021).

14 Epidemiology unit, M. o. H., Sri Lanka. Progress of COVID-19 Immunization as of 07. 12. 2021 2(Epidemiology Unit, Sri Lanka, 2021).

15 Lamikanra, A. et al. Comparability of six different immunoassays measuring SARS-CoV-2 antibodies with neutralizing antibody levels in convalescent plasma: From utility to prediction. Transfusion, doi:10.1111/trf.16600 (2021).

16 Cromer, D. et al. Neutralising antibody titres as predictors of protection against SARS-CoV-2 variants and the impact of boosting: a meta-analysis. Lancet Microbe, doi:10.1016/S2666-5247(21)00267-6 (2021).

17 Tan, C. W. et al. A SARS-CoV-2 surrogate virus neutralization test based on antibody-mediated blockage of ACE2-spike protein-protein interaction. Nature biotechnology 38, 1073–1078, doi:10.1038/s41587-020-0631-z (2020).

18 Tan, C. W. et al. Pan-Sarbecovirus Neutralizing Antibodies in BNT162b2-Immunized SARS-CoV-1 Survivors. The New England journal of medicine 385, 1401–1406, doi:10.1056/NEJMoa2108453 (2021).

19 Chia, W. N. et al. Dynamics of SARS-CoV-2 neutralising antibody responses and duration of immunity: a longitudinal study. Lancet Microbe 2, e240–e249, doi:10.1016/S2666-5247(21)00025-2 (2021).

20 Andrews, N. et al. Vaccine effectiveness and duration of protection of Comirnaty, Vaxzevria and Spikevax against mild and severe COVID-19 in the UK. medRxiv, 2021.2009.2015.21263583, doi:10.1101/2021.09.15.21263583 (2021).

21 Nordström, P. a. B., Marcel and Nordström, Anna,. Effectiveness of Covid-19 Vaccination Against Risk of Symptomatic Infection, Hospitalization, and Death Up to 9 Months: A Swedish Total-Population Cohort Study. (2021).

22 Falsey, A. R. et al. SARS-CoV-2 Neutralization with BNT162b2 Vaccine Dose 3. The New England journal of medicine 385, 1627–1629, doi:10.1056/NEJMc2113468 (2021).

23 Barda, N. et al. Effectiveness of a third dose of the BNT162b2 mRNA COVID-19 vaccine for preventing severe outcomes in Israel: an observational study. Lancet 398, 2093–2100, doi:10.1016/S0140-6736(21)02249-2 (2021).

24 Pulliam, J. R. C. et al. Increased risk of SARS-CoV-2 reinfection associated with emergence of the Omicron variant in South Africa. medRxiv, 2021.2011.2011.21266068, doi:10.1101/2021.11.11.21266068 (2021).

25 Kamaladasa, A. et al. Comparison of two assays to detect IgG antibodies to the receptor binding domain of the SARSCoV2 as a surrogate marker for assessing neutralizing antibodies in COVID-19 patients. Int J Infect Dis, doi:10.1016/j.ijid.2021.06.031 (2021).

26 Heinz, F. X. & Stiasny, K. Distinguishing features of current COVID-19 vaccines: knowns and unknowns of antigen presentation and modes of action. NPJ Vaccines 6, 104, doi:10.1038/s41541-021-00369-6 (2021).

27 Carnell, G. W. et al. SARS-CoV-2 Spike Protein Stabilized in the Closed State Induces Potent Neutralizing Responses. Journal of virology 95, e0020321, doi:10.1128/JVI.00203-21 (2021).

28 Nina Urke Ertesvåg, J. X., Fan Zhou, Sonja Ljostveit, Helene Sandnes, Sarah Lartey Jalloh, Marianne Saevik, Lena Hansen, Anders Madsen, Kristin Mohn, Elisabeth Fjelltveit, Jan Stefan Olofsson, Tiong Kit Tan, Pramila Rijal, Lisa Schimanski, Sir Øyen, Susanna Dunachie, William James, Adam Harding, Heli Harvala, Anni Jämsén, David Roberts, Maria Zambon, PHE Virology group, Karl Broksatd, Oxford Collaborative group, Alain Townsend, Nina Langeland, Bergen COVID-19 Research Group, Rebecca Cox. A Rapid Antibody Screening Haemagglutination Test for Predicting Immunity to Sars CoV-2 Variants of Concern. (2021).

29 Dolgin, E. Is one vaccine dose enough if you’ve had COVID? What the science says. Nature 595, 161–162, doi:10.1038/d41586-021-01609-4 (2021).

