## Supplementary material for "Comparison of the Immunogenicity of five COVID-19 vaccines in Sri Lanka": Detailed methods

**Online methods**

Study participants

Sri Lankan individuals who had received either the AZD1222 (Astrazeneca/Covishield) vaccine, Moderna (mRNA-1273), Sinopharm/BBIBP-CorV, only the first dose of Sputnik/ Gam-COVID-Vac (Sputnik light) or both doses of Sputnik/ Gam-COVID-Vac (Sputnik V), were recruited 12 weeks from receiving the second dose of the vaccine, following informed written consent. In those who received only the first dose of Sputnik/ Gam-COVID-Vac (rAd26-S), blood samples were taken 12 weeks from post-vaccination. The data for individuals who received AZD1222 (12 weeks gap between the two doses)^1^ and the Sinopharm/BBIBP-CorV ^2^ have been previously described. These data on antibody responses to these two vaccines were described in these two studies were used to compare the immunogenicity of these two vaccines with Moderna and Sputnik light and Sputnik V.

In those who received the Moderna, AZD1222 or Gam-COVID-Vac (one or two doses), those who had antibodies to the N protein of the SAR-CoV-2 virus were considered as being infected and were excluded from the analysis. In those who received the Sinopharm/BBIBP-CorV, only those who reported a positive result (either by SARS-CoV-2 specific rapid antigen or PCR) or who reported symptoms of COVID-19, with a family member who was confirmed to have COVID-19, were excluded from the analysis^2^. Details of the N protein assay is described below. The demographic features of the individuals who were included in the analysis of the comparison of the immunogenicity of different vaccines is shown in table 1. As we could not recruit individuals >60 years to study the immunogenicity of the Sputnik vaccines, the immunogenicity for these vaccines was only assessed in individuals <60 years of age.

| Name of the vaccine | Total Number median age (range) | 20 to 39  Number | 40 to 59  Number | >60  Number |
| --- | --- | --- | --- | --- |
| AZD1222  RBD-Abs  ACE2 blocking-Abs | 297  45 years (21 to 81)  69  49 years (24 to 81) | 129  26 | 152  26 | 16  17 |
| Moderna  RBD-Abs  ACE2 blocking-Abs | 224  50 years (22 to 82)  224  50 years (22 to 82) | 48  48 | 132  132 | 44  44 |
| Sinopharm  RBD-Abs  ACE2 blocking-Abs | 203  47 (25 to 72)  110  47 years (25 to 72) | 61  41 | 120  48 | 22  21 |
| Sputnik 1 dose (Sputnik light)  RBD-Abs  ACE2 blocking-Abs | 184  46 (20 to 59)  50  40 (20 to 59) | 45  25 | 139  25 | 0  0 |
| Sputnik 2 doses (Sputnik V)  RBD-Abs  ACE2 blocking-Abs | 127  44 (25 to 66)  127  44 (25 to 66) | 50  50 | 77  77 | 0  0 |

**Table 1: Number of uninfected individuals in each age group who received the different COVID-19 vaccines in Sri Lanka**

The antibody responses in infected individuals who were found to be infected by detection of antibodies to the S protein at the time of recruitment (baseline infected individuals), by detection of antibodies to the N protein or by reporting a COVID-19 infection were analyzed separately.

Ethics approval was obtained from the Ethics Review Committee of University of Sri Jayewardenepura.

Detection of total antibodies to the receptor binding domain (RBD) of SARS-CoV-2

SARS-COV-2 specific total antibody (IgM, IgG and IgA) responses to the RBD were assessed using the Wantai SARS-CoV-2 antibody ELISA (Beijing Wantai Biological Pharmacy Enterprise, China) as previously described according to the manufacturer’s instructions ^3^. The antibody index (an indirect measure of the total antibody levels to the RBD) was calculated by dividing the absorbance of each sample by the cutoff value, according to the manufacturer’s instructions. This assay was shown to have a sensitivity of 98% and was found to be 100% specific when tested using serum samples obtained in 2018, in Sri Lankan individuals ^4^.

Measuring the presence of neutralising antibodies to the SARS-CoV-2 using a surrogate assay

A surrogate virus neutralization test (sVNT) ^5^, which measures the percentage of inhibition of binding of the RBD of the spike protein to recombinant ACE2 (Genscript Biotech, USA) was used to measure the ACE2 blocking antibodies. Inhibition percentage ≥ 25% in a sample was considered as positive for ACE2 blocking antibodies. This assay was found to be 100% specific for measuring ACE2 blocking antibodies in the Sri Lankan population ^6^. The sVNT was only done in a sub cohort of individuals for each of the vaccines. The number of individuals and their age groups used in this assay is shown in table 1.

Assays to determine antibodies to the N protein

Qualitative detection of antibodies to SARS-CoV-2 nucleocapsid (N) antigen was carried out using the Elecsys® Anti-SARS-CoV-2 electrochemiluminescence immunoassay (Cat: 09 203 095 190, Roche Diagnostics, Germany) using the Cobas e 411 analyzer (Roche Diagnostics, Germany). A Cutoff index (COI) ≥1.0 was interpreted as reactive and COI <1.00 was considered non-reactive as per the kit guidelines.

Haemagglutination test (HAT) to detect antibodies to the receptor binding domain (RBD) of VOCs

The HAT was carried out as previously described using the B.1.1.7 (N501Y), B.1.351 (N501Y, E484K, K417N) and B.1.617.2 versions of the IH4-RBD reagents ^7^, which included the relevant amino acid changes introduced by site directed mutagenesis. The assays were carried out and interpreted as previously described and a titre of 1:20 was considered as a positive response ^3,8^. The HAT titration was performed using 7 doubling dilutions of serum from 1:20 to 1:1280, to determine presence of antibodies to the RBD in the different VOCs. The RBD-specific antibody titre for the serum sample was defined by the last well in which the complete absence of “teardrop” formation was observed. A titre of 1:20 was considered as a positive response, as previously determined ^8^.

Statistical analysis

The analysis of data was conducted using the R software (version 4.0.3), R-Studio (version 1.4.1106) and GraphPad PRISM version 8.3. As the data were not normally distributed, differences in the antibody levels measured by different assays for different vaccines were compared using the Mann-Whitney U test (two tailed).

**References:**

1	Jeewandara, C. *et al.* Kinetics of immune responses to the AZD1222/Covishield vaccine with varying dose intervals in Sri Lankan individuals. *medRxiv*, doi:10.1101/2021.10.27.21265561 (2021).

2	Jeewandara, C. *et al.* Persistence of antibody and T cell responses to the Sinopharm/BBIBP-CorV vaccine in Sri Lankan individuals. *medRxiv*, 2021.2010.2014.21265030, doi:10.1101/2021.10.14.21265030 (2021).

3	Jeewandara, C. *et al.* Immune responses to a single dose of the AZD1222/Covishield vaccine in health care workers. *Nat Commun* **12**, 4617, doi:10.1038/s41467-021-24579-7 (2021).

4	Jeewandara, C. *et al.* Transmission dynamics, clinical characteristics and sero-surveillance in the COVID-19 outbreak in a population dense area of Colombo, Sri Lanka April- May 2020. *PloS one* **16**, e0257548, doi:10.1371/journal.pone.0257548 (2021).

5	Tan, C. W. *et al.* A SARS-CoV-2 surrogate virus neutralization test based on antibody-mediated blockage of ACE2-spike protein-protein interaction. *Nature biotechnology* **38**, 1073-1078, doi:10.1038/s41587-020-0631-z (2020).

6	Jeewandara, C. *et al.* SARS-CoV-2 neutralizing antibodies in patients with varying severity of acute COVID-19 illness. *Sci Rep* **11**, 2062, doi:10.1038/s41598-021-81629-2 (2021).

7	Townsend, A. *et al.* A haemagglutination test for rapid detection of antibodies to SARS-CoV-2. *Nature Communications*, 2020.2010.2002.20205831, doi:10.1101/2020.10.02.20205831 (2020).

8	Kamaladasa, A. *et al.* Comparison of two assays to detect IgG antibodies to the receptor binding domain of the SARSCoV2 as a surrogate marker for assessing neutralizing antibodies in COVID-19 patients. *Int J Infect Dis*, doi:10.1016/j.ijid.2021.06.031 (2021).
